# Early Vocal Development in Tuberous Sclerosis Complex

**DOI:** 10.1101/2021.01.06.21249364

**Authors:** Tanjala T. Gipson, Gordon Ramsay, Ellen Ellison, Edina Bene, Helen L. Long, D. Kimbrough Oller

## Abstract

**Objective:** To determine whether entry into the canonical stage, canonical babbling ratios (CBR) and the level of volubility (vocal measures) are delayed in infants with Tuberous Sclerosis Complex (TSC), we completed human coding of their vocalizations at 12 months and compared the results to typically developing infants with no clinical features (TD/NCF).

**Methods:** We randomly selected videos from 40 infants with TSC from the TACERN database. All 78 videos were coded in real-time in AACT (Action Analysis, Coding and Training).

**Results:** Entry into the ***canonical stage*** was delayed in the great majority of the infants with TSC. The ***CBR*** for the TD/NCF infants was significantly higher than for the infants with TSC (TD/NCF mean = .346, SE = .19; TSC mean = .117, SE = .023). ***Volubility*** level in infants with TSC was less than half that of TD/NCF infants (TD/NCF mean = 9.82, SE = 5.78; TSC mean = 3.99, SE = 2.16). CBR and volubility were also lower in TSC infants than in TD/NCF infants recorded all-day at home.

**Conclusions:** Entry into the canonical stage was delayed, while canonical babbling ratios and volubility were low in infants with TSC. Assessing prediction of neurodevelopmental outcome using these vocal measures in combination with non-vocal measures will be the focus of planned studies in this high-risk population.

## Introduction

Tuberous Sclerosis Complex (TSC) is a genetic disease characterized by non-cancerous tumors in the brain, kidneys and heart [1]. TSC can cause developmental problems [2, 3] and up to 50% have autism spectrum disorder (ASD) [4]. Social communication deficits are similar to non-syndromic ASD (nsASD), and early intervention is recommended pre-symptomatically [5]. However, ASD in TSD is typically diagnosed at a median age of 7.8 years [6]. Additionally, only 28% of 510 individuals with TSC showed normal language development [7]; however, detailed studies of language impairments and ability are lacking [2]. Language laterality abnormalities have been identified in patients with TSC. MEG revealed left-hemisphere language dominance in 73.3% and a bilateral language pattern in 26.7% of 15 patients with TSC and epilepsy. Left-hemisphere dominance is typically present in 94-96%. There were also more cortical tubers in language regions of individuals with a bilateral pattern [8].

In TSC, epilepsy occurs in 90% [9], and the neurodevelopmental impact has been assessed. Infantile spasms and early onset epilepsy were noted to be risk factors for ASD [9]. ASD by 36 months was also predicted by seizure onset prior to 6 months, higher seizure frequency, and associated with language scores at 12 months from the Mullen Scales of Early Learning (MSEL), the Vineland Adaptive Behavior Scales 2^nd^ edition (VABS-II), and the Preschool Language Scales, 5^th^ edition (PLS-5) [10].

Among 130 children from TACERN, seizures predicted worse performance on all developmental assessments, and children with infantile spasms had lower scores. Increased seizure frequency also predicted poor developmental outcome [11]. Intellectual disability was linked to severe and early-onset epilepsy in the first 2 years in a study of 88 children with TSC as compared to their siblings. [12].

Preventing epilepsy, the goal of recent trials using the TSC-targeted medications everolimus and vigabatrin, is a logical next step given these risk factors [13-16] [17]. Thus far, improved neurodevelopmental outcomes have been observed among infants with TSC who received vigabatrin prior to seizure onset [18].

Unfortunately, we are not able to reliably predict which infants are at highest risk for poor neurodevelopmental outcome prior to intervention [4]. Improving our ability to predict neurodevelopmental outcomes in TSC by assessing vocal development in combination with other existing measures is the goal of our current work.

### Rationale for studying vocal development in TSC

Identification of early, accessible, generalizable biomarkers for ASD and language disorders in TSC would help guide the *timing* and *type* of interventions. The aim of the current work, focused on infants with TSC at 12 months, is to assess the utility of objectively analyzing infant vocalizations as one of several potential predictors. In prior work, an infrastructural theory of language origins with important focus on precursors to speech, or “protophones”, has been developed [19]. These initial infant vocalizations, which exclude cries and laughs, reveal foundations for speech because the capacity for voluntary and flexible vocalization is a requirement for all aspects of vocal language [20]. During the first few months of life, the *precanonical* protophones include vocants (vowel-like sounds), squeals, and growls [21, 22]. *Volubility* can be measured as the total number of protophones or individual syllables produced by an infant. Research shows that human infants tend to produce very high volubility, several protophones per minute across the entire first year [23-25].

*Canonical babbling* (CB) develops during the 2^nd^ half of the first year and includes well-formed ‘syllabic’ consonant-vowel combinations that are more speech-like than the early protophones [26, 27] [28, 29]. In canonical syllables phonation co-occurs with supraglottal movements [30] yielding syllables such as ‘ba’, ‘da’ and ‘na’ [31]. The variable pronunciation of the early protophones and canonical syllables [32, 33] resembles the pattern of variability observed in early arm and hand movements, which become increasingly purposeful across time [34]. There has been major emphasis recently on the study of canonical babbling as a precursor to speech as indicated in a meta-analysis of many studies by Morgan & Wren [35].

Canonical babbling onset (usually around 7 months and overwhelmingly by 10 months in typically developing infants) is a necessary foundation for speech [20, 36] and predicts speech disorders; significant delays occur in onset of canonical babbling in Williams Syndrome [37], deafness [38, 39] [40], Down Syndrome [41], cleft palate [42, 43], cerebral palsy [44], Fragile X syndrome [45], childhood apraxia of speech [46], and autism [47]. Other disorders related to language development have also been indicated as involving significant deficiencies in canonical babbling [48-50].

A measure often used to assess canonical stage status in infants is the canonical babbling ratio (CBR), the number of canonical syllables in a recorded sample divided by the total number of syllables. Children in the canonical stage are generally expected to show a minimum CBR of .15 [41]. There has also been considerable research on the validity of the CBR and its role in prediction of disorders [51] [52] [53].

There are several reports of low volubility in infants with autism [47], Fragile X syndrome, [45], and childhood apraxia of speech [46]. Thus, both low volubility and delays in the development of canonical babbling have been shown to be involved in a host of disorders of language. Yet neither volubility nor canonical babbling have been assessed to our knowledge in infants with TSC.

Data about infant vocalizations can be acquired successfully using laboratory recordings, parent interviews, family videos, and in-the-home all-day recordings using the LENA device worn in a vest by infants [54]. Using all-day recordings, automated vocal analysis of 232 children with 1,486 all-day recordings reliably distinguished infants with typical development, language delay, and autism spectrum disorder [55]. Other research with LENA automated analysis of all-day recordings has also provided significant evidence on the predictiveness of its measures for language and cognition ten years later [56]. Other work with all-day recordings has shown important relations between the measures and language disorders, especially in autism [57-59]. However, this approach is applicable to groups but not appropriate for comparisons across individual infants, and thus not a sound basis for screening, diagnosis, or simply for projections regarding language development in individual children. The more demanding methods of human listening and judgments provide the gold standards for analyzing the vocalizations of individual infants and provide the appropriate starting point for evaluations of predictors of language outcome in TSC. Here, we present preliminary findings using the analysis of infant vocalizations in TSC based on human coding for recorded vocal samples. Our aim is in part to develop necessary tools to contribute to early identification of autism spectrum disorder and language disorders in TSC using this technique. The vocal analysis approach will be supplemented by assessment of additional potential predictors in future efforts from our collaboration.

The age we selected to evaluate canonical babbling is ∼12 months, in part because by this age a wide body of evidence shows that typically developing infants should be in the canonical stage [22, 60-62]. The achievement of the canonical stage before 12 months is robust in infants who do not develop language-related disorders—for example even infants born prematurely, without significant perinatal problems, develop canonical babbling on the normal schedule [63], as do infants of low socio-economic status [64, 65]. However, given that many infants with speech-related disorders show CB onset beyond 12 months and/or low CB ratios (see citations above), this age provides an appropriate target for evaluation in TSC, where delays of vocal development seem distinctly possible. Still, no prior research to our knowledge has evaluated canonical babbling in infants with TSC. The work is intended to contribute to an understanding of predictors for neurodevelopmental language outcomes in infants with TSC in hopes of helping to provide earlier identification.

### Objectives

The first objective of this study is to develop a set of human coded vocalizations based on laboratory-based recordings for 40 infants with TSC at 12 months. There has never been any such dataset to our knowledge. Based on these data we will:

1. calculate CBRs and compare these ratios to those of infants at 12 months based the original data available to us from coding of infant laboratory recordings in a published paper on canonical babbling [66] of typically-developing infants with no known clinical features (TD/NCF);
2. compare the onset of CB in the TSC infants in laboratory recordings (gauged from the CBRs in accord with a criterion from the existing literature) with those of the TD/NCF infants, determining whether infants with TSC may be delayed in onset;
3. compare volubility (the amount of vocalization independent of CB) in infants with TSC in laboratory recordings with volubility data from the laboratory recordings of TD/NCF infants [66].

Although the existing literature is unambiguous in the conclusion that TD/NCF infants should show CBRs exceeding .15 and thus indicating canonical stage status by 12 months in accord with the standard model (see citations above), the methodology for determining both CBR and canonical stage status has been notably variable across studies (see Molemans, et al., 2012). Presumably as a result of these methodological differences, substantial variations have been reported in CBR values and in ages at which infants have been reported to reach the canonical stage (again see Molemans, et al. 2012). Further, volubility in TD/NCF infants has been reported to differ widely across studies; again, it appears methodological differences across studies may have accounted for the different outcomes. Thus, there remain important open questions about the possible effects of methodological variations on all the matters at hand: CBR, canonical stage status, and volubility. Of course, the optimal comparison for the present paper would be methodologically matched for the two groups: since the TSC infant data were recorded at 12 months in the particular test situations of the TACERN study (ESCS and AOSI), it would be optimal to compare with data from TD infants recorded in the very same test situations. We do not have access to such data, but the available data based on laboratory recordings of TD infants appear to provide the currently optimal comparison set.

However, to help put the data on infants with TSC in additional perspective, recognizing that different methodologies may yield substantially different quantitative outcomes, we can provide an additional comparison of the TSC samples with data recently acquired from all-day recordings of TD/NCF infants. The all-day recording method is rapidly providing new perspectives on vocal development (see citations above). While automated analysis offers hope for the future, the point of comparison needed for the TSC data is one where the all-day recordings are studied by random sampling of segments that are then human coded. This method offers, presumably, a more representative view of infant vocal capabilities than views provided by other common methods, namely laboratory recordings, test-based recordings, or short in-the-home recordings. The study from which the data available to us from all-day recordings have been acquired is ongoing, see Oller et al. (2020) [67]. These data allow us to:

1. provide a comparison of the TSC outcomes with data on development of CBR and canonical stage status in 12-month old TD/NCF infants who have been studied with all-day recordings, and to
2. assess the volubility produced by infants with TSC compared with data from the same all-day recordings of TD/NCF infants.

We hypothesize based on comparisons with existing literature that:

1. CBRs will be lower in TSC infants in laboratory recordings than CBRs reported for typically developing infants in laboratory recordings,
2. CB stage status determined by the literature’s CBR criterion will be delayed among infants with TSC at 12 months, and that
3. the level of volubility will be lower in the TSC infants in laboratory recordings than the level reported in the existing literature based on laboratory recordings iTD/NCF infants.

Based on comparisons with the all-day recordings, we predict that:

1. CBRs will be lower in the TSC infants than in the all-day recordings of TD/NCF infants with no clinical features, and that
2. the level of volubility will be lower in the TSC infants than levels found for the TD/NCF infants based on the all-day recordings.

The long-term goal of this work is to assess behavioral predictors of neurodevelopmental significance in TSC, and here we will ultimately include measures of vocal, gestural, affective, and social responsivity development. Assessing these factors is important because identifying infants at risk for autism and language disorders earlier would allow us to reliably test the ability of available neurobiologically targeted medications to alter negative outcomes. Our short-term goals are more modest, focused on vocal development, as expressed in the five predictions above.

## Methods

### Participants

#### Data acquired through the TACERN study (Source of data used for infants with TSC)

Video recorded data for the current study were obtained and made available for human coding from the TSC Autism Center of Excellence Research Network (TACERN). TACERN is comprised of 5 children’s hospitals across the US, which conducted The Early Biomarkers of Autism in Infants with TSC study which included 130 infants [11]. (See inclusion and exclusion criteria in Supplement [68].)

#### Data collection in the TACERN work, Early Biomarkers of Autism in Infants with TSC, the longitudinal study

Developmental and adaptive assessments were completed at ages 3, 6, 9, 12, 18, 24 and 36 months in the TACERN study. For the current study, the data of most relevance are the video-recorded materials from the ESCS and the AOSI at 12 months (see below). See supplement for details about assessments.

### Study Procedures

#### Early Vocal Communication in TSC (current study)

After obtaining a data sharing agreement and IRB approval by the University of Tennessee Health Sciences Center and Cincinnati Children’s Hospital, we received the complete data set from TACERN’s Early Biomarkers of Autism in Infants with TSC study (N=130) for the current study focused on audio-video recordings obtained at 12 months during evaluations using the ESCS and/or the AOSI. We randomly selected 40 infants for initial analysis on vocal development. Of the 40 infants, 38 had 2 recordings, and 2 infants had 1; therefore, 78 recordings were available for the analysis. The mean duration of the recordings for each of the tests was 15.1 minutes, with average duration across the two tests for each infant in the final sample being 30.4 minutes. An analysis of CBR in the recordings of the ESCS compared with the AOSI revealed that the number of syllables available for analysis was nearly doubled by collapsing the two samples for each child (two of the 38 had one of the two tests), so the data for the two recordings were collapsed for the analyses below.

#### Coding of Infant Vocalizations

Training for coding: The last author of the present paper trained the first and third authors using the coding training protocol of Origin of Language Laboratories (OLL) [66], and good coding agreement was established (see Supplement).

## Results

Figure 1 presents the data on the five objectives.

**Figure 1:**
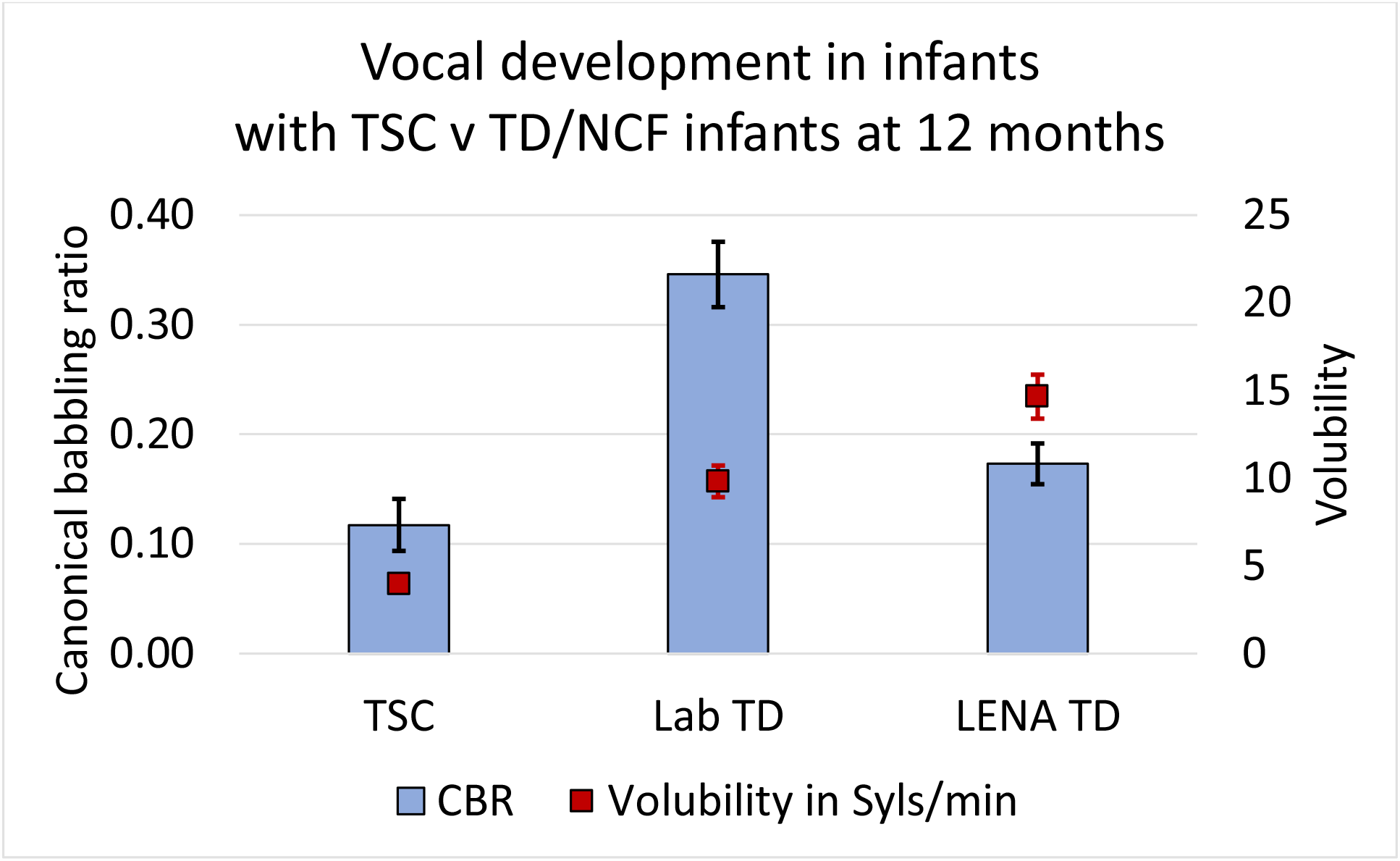
Both canonical babbling (reflected in the canonical babbling ratio or CBR) and volubility (reflected in syllables per minute of speech-like vocalization) were lower in infants with TSC than in TD/NCF infants based on two comparison samples at 12 months. The optimal comparison, we think, is between data based on laboratory recordings of 38 infants with TSC and 41 TD/NCF infants also based on laboratory recordings. A secondary comparison can be made between the laboratory recordings of 38 infants with TSC and all-day (LENA) recordings on 39 TD/NCF infants. (See Supplement for details about the comparison samples.) Note also that although the two comparison samples yielded higher values than the TSC sample, they were also considerably different from each other, with CBR being much higher in the laboratory than in the all-day recordings, while volubility was much higher in the all-day (LENA) recordings. Standard errors are displayed, although for the volubility of the infants with TSC in laboratory recordings, the SE was so low (.35), it is not visible in the scaling of the data in Figure 1.

### 1. Canonical babbling ratios (CBRs) in infants with TSC in the perspective of the existing literature in vocal development

To our knowledge the existing literature for TD/NCF infants at 12 months provides only one well-documented sample reporting CBR at 12 months, using very similar human coding procedures to those of the current research (Oller, et al. 1997). In addition, the sample in question evaluated 41 infants, a number comparable to that of the present work (38). The CBR for the TD/NCF infants was dramatically higher than for the infants with TSC (TD/NCF mean = .346, SE = .19; TSC mean = .117, SE = .023). By t-test, the CBRs were very significantly different with a very large effect size (*p* < .0001, *d* = 1.34, 95% CI [0.93, 1.75]).

### 2. Canonical stage onset status in infants with TSC in the perspective of the existing literature in vocal development

The TSC infants can be classified with regard to canonical stage status based on the criterion of .15 CBR. Only 26% of the infants with TSC had CBRs exceeding that value, with only 13% showing CBRs as high as the mean for the TD/NCF infants in the published comparison study. By contrast, >90% of the TD/NCF infants in the comparison study had CBRs higher than .15 at 12 months. In addition, another publication based on similar methods [51] (while not providing CBRs for each infant, nor mean values at 12 months) indicated that all of the 40 TD/NCF infants in their longitudinal study met the canonical stage criterion of .15 in recordings obtained at 12 months.

### 3. Volubility in infants with TSC in the perspective of the existing literature in vocal development

Our measure of volubility for the TSC infants is in terms of syllables, both canonical and non-canonical. The infants with TSC showed volubility levels less than half that of the estimated levels in syllables per minute for TD/NCF infants based on laboratory recordings in the Oller et al. (1997) study (TD/NCF mean = 9.82, SE = 5.78; TSC mean = 3.99, SE = 2.16). As in the case of CBR, the volubility differences by t-test were very significantly different and showed a very large effect size (*p* < .0001, *d* = 1.34, 95% CI [0.93, 1.75]).

### 4. CBRs of infants with TSC compared with data based on all-day recordings of TD/NCF infants

The comparison of CBRs in infants with TSC and TD/NCF infants recorded all-day from the ongoing study from the OLL in collaboration with Emory University, the Marcus Autism Center as part of an NIH Autism Center of Excellence, also suggested low CBRs for the TSC infants. In this work, 39 TD/NCF infants provided all-day recordings at 12 months. (TD/NCF mean = .173, SE = .019; TSC mean = .117, SE = .023). A t-test of the data from the TSC infants and the TD/NCF infants yielded a statistically significant difference (*p* < .04), with a moderate effect size (*d* = 0.42, 95% CI [0.04-0.80]). However, with Bonferroni correction for the fact that CBR is tested twice here (once for TSC vs TD/NCF laboratory recordings and once of TSC vs. TD/NCF all-day recordings), this t level falls just short of the adjusted alpha value of .025. The data from 59% of the TD/NCF infants met the .15 CBR criterion for onset of the canonical babbling stage, while only 26% if the infants with TSC met the criterion.

### 5. Volubility of infants with TSC compared with data from all-day recordings of TD/NCF infants

The quantitatively largest difference between the infants with TSC and TD/NCF infants concerned volubility when comparing with the data from the all-day recordings. The rate of vocalization in syllables per minute was more than three times higher in the TD/NCF infants based on all-day recordings (TD/NCF mean = 14.65, *SE* = 1.25; TSC mean = 3.99, *SE* = 2.16). By t-test, the volubility levels were very significantly different, with a very large effect size (*p* < .0001, *d* = 1.86, 95% CI [1.41, 2.31]).

## Discussion

Given the high prevalence of ASD in TSC, along with significant language delays in infants with TSC, early identification of language-related delays is imperative in TSC. In the present study, we examined for the first time the early vocalizations of infants with TSC. Canonical babbling specifically is indicative of developmental progression, and infants not reaching the canonical babbling stage by age 10 months are considered at risk for a variety of disorders including those that affect language development. Examination of canonical babbling in infants with TSC in the present study established the feasibility of acquiring an assessment of vocal development in TSC from stored, audio-video data.

The results summarized in Figure 1 show that both canonical babbling and volubility were lower in infants with TSC than in two similarly-sized comparison samples. Although there were some infants with TSC who showed typical rates of canonical babbling, the great majority displayed low rates when compared to TD infants without clinical features. The infants with TSC were much less likely than the TD/NCF infants to be classifiable as being in the canonical stage based on the CBRs. Even more strikingly, the infants with TSC showed very low volubility. They simply did not vocalize as often as one would expect for TD/NCF infants based on the comparison samples and the existing literature on vocal development.

We also highlight the large differences between outcomes for TD/NCF infants based on the two comparison studies. In fact, CBR was significantly different for the two comparison studies (*p* < .0001) with a large effect size (*d* = 1.09). These differences confirm the observations of Molemans et al. (2012) which suggested important differences in outcomes for CBR based on different methods following a review of studies on this topic. The comparison of the two TD/NCF groups also adds evidence showing that all-day recordings — which are becoming an increasing common method of research in vocal development — may yield important evidence, contrasting with that from the more common methods of the past.

Of similar interest are the significant differences between the two TD/NCF studies on volubility (*p* < .01) which corresponded to a moderate size for volubility (*d* = .70). Especially surprising is the fact that the differences between the two comparison studies go in opposite directions for CBR and volubility. Specifically, CBR was higher for the laboratory recordings and volubility was higher for the all-day recordings. We have no explanation at present for these differences.

Determining and acknowledging such differences, however, within studies of vocal development in TD/NCF infants is clearly important because it illustrates the complexity of the task ahead for those of us who seek to use infant vocal development to gauge possible anomalies in infants with disorders such as TSC. While the outcomes suggest infants with TSC have low volubility and CBR as a group, we cannot rule out at present that methodological characteristics of the TSC data may have contributed to the apparent effects, especially given that there were significant differences between the two TD/NCF comparison groups. The most salient characteristic of the TSC data that contrasts with the comparison studies is that during these laboratory recordings, the infants (while sitting next to the mothers) were interacting with an examiner who conducted the tests (ESCS and AOSI), while the comparison studies (both in the laboratory and in the home for all-day recordings) were conducted with the parent as the primary interactor with the infant. In the comparison studies, any non-relative in the laboratory recordings was at least slightly familiar to the infants because they had been present in several prior recordings of the longitudinal research study. Thus, the ubiquitous presence of a stranger (test examiner) in the TSC recordings might be thought to have inhibited both volubility and CBR. In the future, we aim to conduct a study that should disambiguate this interpretation using comparison recordings in test circumstances of either the AOSI or the ESCS or both with TD/NCF infants at 12 months.

The sample size of our study was relatively small, but a larger sample size will be possible in research in the near future. Additionally, there were fewer TSC infants (16%) who had an ASD diagnosis than would be expected based on the reported prevalence of 50% of infants with ASD in TSC. Perhaps notably the infants who did have an autism diagnosis showed practically identical CBR and volubility rates to those of the entire TSC group. Consequently, it may be concluded in very preliminary fashion that infants with TSC may present with low CBRs and low volubility independent of ASD; however, a larger sample size with more infants who later receive an ASD diagnosis is necessary to support this claim.

## Data Availability

Portions of these data will be made available on the OSF following journal acceptance.

## Acknowledgements

Tanjala T. Gipson, MD designed and conceptualized study; acquired, analyzed, and interpreted the data and both drafted and revised the manuscript for intellectual content. Gordon Ramsay, PhD acquired the data and revised the manuscript for intellectual content. Ellen Ellison analyzed the data and reviewed the manuscript. Edina Bene, PhD acquired, analyzed, and interpreted the data and revised the manuscript for intellectual content. Helen L. Long, PhD interpreted the data and revised the manuscript for intellectual content. D. Kimbrough Oller, PhD designed and conceptualized study; acquired, analyzed, and interpreted the data and revised the manuscript for intellectual content.

The authors gratefully acknowledge the TACERN consortium for data sharing.

## Supplement

### Inclusion/exclusion criteria TACERN Early Biomarkers of Autism in Infants with TSC study

TACERN is comprised of 5 children’s hospitals across the US. The Early Biomarkers of Autism in Infants with TSC study which included 130 infants [11] was conducted by this network with the following inclusion/exclusion criteria.

#### Inclusion criteria

1. Meets genetic or clinical diagnostic criteria for TSC, the latter based on current recommendations for diagnostic evaluation, such as physical exam, neuroimaging and/or echocardiogram.
2. Age criteria: 3-12 months of age at time of enrollment.

#### Exclusion criteria

1. Prematurity, defined as gestational age <36 weeks at time of delivery
2. Has taken an investigational drug as part of another research study, within 30 days prior to study enrollment
3. Is taking an mTOR inhibitor such as rapamycin, sirolimus or everolimus (other than topical formulations) at the time of study enrollment
4. Subependymal giant cell astrocytoma requiring medical or surgical treatment at the time of study enrollment
5. History of epilepsy surgery at the time of study enrollment
6. Contraindications to MRI scanning, such as metal implants/non-compatible medical devices or medical condition

## Supplement

### Developmental assessments completed during The Early Biomarkers of Autism in Infants with TSC study

Early Social Communication Scales (ESCS) was completed at 12 months with video [69]. To assess for autism, the Autism Observation Scale for Infants (AOSI) [70] was completed at age 12 months with video. The Mullen Scales of Early Learning [71] was completed at each visit as well as the Vineland Adaptive Behavior Scales, 2^nd^ edition, specifically the Survey Interview, which was completed at ages 6, 12, 18, 24 and 36 months [72].The Mullen Scales of Early Learning [71] was completed at each visit as well as the Vineland Adaptive Behavior Scales, 2^nd^ edition, specifically the Survey Interview, which was completed at ages 6, 12, 18, 24 and 36 months [72]. The Preschool Language Scale-5^th^ edition was completed at each visit [73]. The Autism Diagnostic Observation Schedule-2 [74] and Autism Diagnostic Interview-Revised [75] was completed at ages 24 and 36 months [73].

### Coding training for the current study

Specifically, training began with a two-hour introduction using audio and audio-video examples of infant vocalizations. The last author’s (Oller) career spans decades (with publications as early as 1976 [26]) and has been focused on infant vocal development and establishing descriptive protocols for prelinguistic sounds. AACT (Action Analysis, Coding and Training [76]), a software coding and analysis system under development since the 1980’s in and associated with the Origin of Language Laboratories (OLL), was also introduced in this initial session. The first and third authors were trained to use this system for real-time and repeat-listening coding in AACT, where coders can view video simultaneously with synchronized audio displays of both waveforms and spectrograms. The system is described in detail in the Supplementary Material to Oller et al. [77]. The next step in training was completion of a series of coding assignments using a set of coding-training recordings used routinely in OLL training. They had been previously coded by at least 30 trainees. During the training for the TSC study, these samples were coded both alone and in collaboration with a senior coder (another member of the OLL coding team or a PhD student who had completed training previously). Over the course of 6-8 weeks, several cycles of coding and result reviews were completed, with weekly group and individual meetings as needed for results in need of correction. Readiness for coding study data was established when the coder fell within 10% of the cry and protophone counts for a gold standard set of five sessions coded by the last author.

Coding categories and procedure: All 78 TSC videos were coded in real-time, using the same procedure that is routinely used in OLL “Phase 2 coding” (see Oller et al., 2019) [25]. Each video was reviewed in real-time in AACT. Coding of infant vocalizations focused on protophones, the presumed precursors to speech, which include squeals, growls, vowel-like sounds (vocants), and whispers. In Phase 2 coding the only task was to produce a single keystroke for each canonical syllable and a different one for each non-canonical syllable as they occurred in the protophones in real-time. ∼ 13% of coded infant syllables were determined to be canonical with the remainder noncanonical. Because only protophones were evaluated in Phase 2 coding, the infants’ cries, laughs, and effort grunts were not included in any of the analyses. The coding made possible the computation of the canonical babbling ratio (CBR), the quotient of the number of canonical syllables and the number of all syllables counted in each sample. This is the primary measure to be compared across the infants with TSC and infants with no known risk of disorders of communication.

Although the current analysis does not include adult vocalizations, all sentences or phrases voiced by an adult during the recordings were also coded as either directed to another adult (7% of coded adult utterances), intended for an infant but spoken in an adult register (3% of coded adult utterances), or intended for an infant and spoken in an infant register (89% of coded adult utterances). This pattern is consistent with the fact that a standardized test was being administered, where the adult vocalizations overwhelmingly consisted of prompts or instructions by the adult tester speaking to the infant.

Prior research has supported naturalistic coding (often real-time coding as conducted in the present work) as being theoretically justified and empirically consistent with outcomes from the much more time-consuming method of repeat-observation coding [32, 45, 47]. Syllables are defined in this approach as rhythmic prominences within a vocal breath group. Canonical babbling stage and CBRs were assessed for all except 2 infants with TSC, who were excluded due to low syllable counts. Primary analyses were thus conducted on 38 infants with TSC. Canonical babbling stage judgments and CBRs were compared to controls from the OLL archives.

#### Coding agreement for the current study

All the recordings from the 38 infants were coded in random order by a “primary” coder (EE) trained in the OLL approach with interrater agreement established on a subset of the videos. Comparison of the number of syllables present and CBRs in the AOSI videos as compared to the ESCS videos revealed no significant differences.

Consequently, it was deemed reasonable to select only one of the test types randomly (AOSI) to use in the agreement analyses. A subset of 10 of the 78 recordings for the AOSI test were randomly selected and re-coded by the primary coder in a new random order, several weeks after the original coding and completely blind to the original coding results. An additional “agreement” coding was conducted by a member of the OLL coding team, completely blind to the outcomes of the primary coding at either the original or the re-coded points. A simple correlational analysis showed very high agreement on all the relevant issues: all *r* values (n = 10) exceeded .9 (primary v. agreement coder on number of syllables *r* = .91, on CBR, the key measure, *r* = .99; for the primary coder’s original coding v. her recoding on number of syllables *r* = .99, on CBR, *r* = .96). The mean number of syllables and CBR for the 10 samples for the primary coder on the original coding were 49.3 and .18, for the re-coding 53.2 and .15. For the agreement coder the values were 42.9 and .13. The discrepancies in means are typical of differences found in agreement evaluations using this coding scheme for other coders in the OLL.

Although the current analysis does not include adult vocalizations, all sentences or phrases voiced by an adult during the recordings were also coded as either directed to another adult (7% of coded adult utterances), intended for an infant but spoken in an adult register (3% of coded adult utterances), or intended for an infant and spoken in an infant register (89% of coded adult utterances). This pattern is consistent with the fact that a standardized test was being administered, where the adult vocalizations overwhelmingly consisted of prompts or instructions by the adult tester speaking to the infant.

#### Comparison data from the existing literature on canonical babbling in typically developing infants (Source of data on TD/NCF infants based on laboratory recordings)

As reviewed above, four decades of research are now available on the phenomenon of canonical babbling [20, 22, 46]. That work has illustrated that canonical babbling onset typically occurs by 10 months, with full mastery by 12 months, and that late onset is a risk indicator for delayed or disordered language development (see prior citations). The criterion level for CBR of .15 has been treated as a standard for human-coded samples to indicate canonical stage status, and existing data provide comparative standards for TD/NCF infants at the 12-month age [66]. The original data from the cited study for TD/NCF infants are available to us within the Origin of Language Laboratory (OLL) archives, which is directed by the last author. The data are based on the compilation of several longitudinal studies where audio recordings were made throughout the first year of life. 41 TD/NCF infants provided recordings with human-coded data on the first 70 protophones (speech-like vocalizations) at 12 months. The laboratory at the University of Miami that had collected the data was under the direction of the last author during the data collection and followed protocols of coding that included the same differentiation between canonical and non-canonical syllables as well as the same method of computing CBR as in the present study. While the present study is based on real-time coding of audio-video samples of test situations (ESCS, AOSI), the Miami data involved repeat-observation coding of audio-only samples of interaction with infant parent and laboratory personnel. For details on the methods and participants in the Miami data see Oller et al. (1997) [66].

#### Comparison data acquired from coding of all-day recordings on TD/NCF infants

Relevant data, already having been coded in prior work [25] [67], on infants with no clinical features and no known disorders were also obtained from the archives of OLL at the University of Memphis and utilized as a comparison sample. These data were recorded in Atlanta at the Marcus Autism Center (MAC) and Emory University School of Medicine, as part of a long-term longitudinal study of autism in infancy funded by NIH as an Autism Center of Excellence (NIH P50 MH100029). The longitudinal study is on-going in a collaboration between the MAC and the University of Memphis OLL. There were 39 infants with no clinical features for whom all-day audio-only recordings were available from this collaboration. The recordings were obtained with the LENA device, a small battery-powered recorder worn in an infant vest [78]. Human coding had been conducted in the OLL on 8 five-minute segments selected on the basis of vocal activity level determined after a review of 21 randomly selected segments for each of the all-day recordings. The coding had been conducted using the same procedures as for the current work with the TACERN infants. For further description of the infants and the coding procedures see Oller et al. (2020) [67], especially the Supplementary Information.

## References

1. Kwiatkowski, D.J., Tuberous sclerosis, in Genetic Diseases of the Kidney. 2009, Academic Press. p. 527–542.

2. Prather, P. and P.J. de Vries, Behavioral and cognitive aspects of tuberous sclerosis complex. J Child Neurol, 2004. 19(9): p. 666–74.

3. Hunt, A., Development, behaviour and seizures in 300 cases of tuberous sclerosis. J Intellect Disabil Res, 1993. 37 (Pt 1): p. 41–51.

4. Curatolo, P., R. Moavero, and P.J. de Vries, Neurological and neuropsychiatric aspects of tuberous sclerosis complex. Lancet Neurol, 2015. 14(7): p. 733–45.

5. Jeste, S.S., et al., Symptom profiles of autism spectrum disorder in tuberous sclerosis complex. Neurology, 2016. 87(8): p. 766–72.

6. Kingswood, J.C., et al., TuberOus SClerosis registry to increase disease Awareness (TOSCA) - baseline data on 2093 patients. Orphanet J Rare Dis, 2017. 12(1): p. 2.

7. de Vries, P.J., & Bolton, P. F., Tuberous sclerosis, in Outcomes in Neurodevelopmental and Genetic Disorder, P.H. Udwin, Editor. 2002, Cambridge University Press: Cambridge, UK p. 272–298.

8. Gallagher, A., et al., Decreased language laterality in tuberous sclerosis complex: a relationship between language dominance and tuber location as well as history of epilepsy. Epilepsy Behav, 2012. 25(1): p. 36–41.

9. Webb, D.W., A.E. Fryer, and J.P. Osborne, Morbidity associated with tuberous sclerosis: a population study. Dev Med Child Neurol, 1996. 38(2): p. 146–55.

10. Schoenberger, A., et al., Language predictors of autism spectrum disorder in young children with tuberous sclerosis complex. Epilepsy Behav, 2020. 103(Pt A): p. 106844.

11. Capal, J.K., et al., Influence of seizures on early development in tuberous sclerosis complex. Epilepsy Behav, 2017. 70(Pt A): p. 245–252.

12. Tye, C., et al., Long-term cognitive outcomes in tuberous sclerosis complex. Dev Med Child Neurol, 2020. 62(3): p. 322–329.

13. French, J.A., et al., Adjunctive everolimus therapy for treatment-resistant focal-onset seizures associated with tuberous sclerosis (EXIST-3): a phase 3, randomised, double-blind, placebo-controlled study. Lancet, 2016. 388(10056): p. 2153–2163.

14. Krueger, D.A., et al., Everolimus for subependymal giant-cell astrocytomas in tuberous sclerosis. N Engl J Med, 2010. 363(19): p. 1801–11.

15. Franz, D.N., et al., Long-Term Use of Everolimus in Patients with Tuberous Sclerosis Complex: Final Results from the EXIST-1 Study. PLoS One, 2016. 11(6): p. e0158476.

16. Bissler, J.J., et al., Everolimus for renal angiomyolipoma in patients with tuberous sclerosis complex or sporadic lymphangioleiomyomatosis: extension of a randomized controlled trial. Nephrol Dial Transplant, 2016. 31(1): p. 111–9.

17. Wheless, J.W., et al., Treatment of pediatric epilepsy: European expert opinion, 2007. Epileptic Disord, 2007. 9(4): p. 353–412.

18. Jóźwiak, S., et al., Antiepileptic treatment before the onset of seizures reduces epilepsy severity and risk of mental retardation in infants with tuberous sclerosis complex. Eur J Paediatr Neurol, 2011. 15(5): p. 424–31.

19. Oller, D.K., The Emergence of the Speech Capacity. Vol. 428. 2000, Mahwah, NJ: Lawrence Erlbaum Associates.

20. Oller, D.K., The emergence of the sounds of speech in infancy., in Child phonology., J.K. G. Yeni-Komshian, and C. Ferguson, Editor. 1980, Academic Press: New York. p. 93–112.

21. Holmgren, K., On the phonetics of infant vocalization., in Precursors of early speech, B.L.a.R. Zetterstrom, Editor. 1986, Stockton Press: New York. p. 51–63.

22. Stark, R.E., Stages of speech development in the first year of life, in Child Phonology, J.K. G. Yeni-Komshian, and C. Ferguson, Editor. 1980, Academic Press: New York. p. 73–90.

23. Iyer, S.N., et al., Volubility of the human infant: Effects of parental interaction (or lack of it). Clin Linguist Phon, 2016. 30(6): p. 470–88.

24. Locke, J.L., & Pearson, D., Vocal learning and the emergence of the phonological capacity., in Phonological Development: Models, Research, Implications, L.M. C. Ferguson, & C. Stoel-Gammon, Editor. 1992, York: Timonium, MD. p. 91–130.

25. Oller, D.K., et al., Preterm and full term infant vocalization and the origin of language. Sci Rep, 2019. 9(1): p. 14734.

26. Oller, D.K., et al., Infant babbling and speech. Journal of Child Language, 1976. 3: p. 1–11.

27. Stark, R.E., Infant vocalization: A comprehensive view. Infant Medical Health Journal, 1981. 2(2): p. 118–128.

28. Kent, R.D., & Bauer, H., Vocalizations of one year olds. Journal of Child Language, 1985. 12: p. 491–526.

29. Koopmans-van Beinum, F.J., Clement, C. J., & van den Dikkenberg-Pot, I., Babbling and the lack of auditory speech perception: A matter of coordination? Developmental Science, 2001. 4(1): p. 3–14.

30. Oller, D.K. and M.P. Lynch, Infant vocalizations and innovations in infraphonology: Toward a broader theory of development and disorders, in Phonological Development: Models, Research, Implications, C. Ferguson, L. Menn, and C. Stoel-Gammon, Editors. 1992, York Press, Inc.: Parkton, MD. p. 509–538.

31. Locke, J.L., Phonological acquisition and change.. 1983, New York: Academic Press.

32. Ramsdell, H.L., et al., Identification of prelinguistic phonological categories. Journal of Speech Language and Hearing Research, 2012. 55: p. 1626–1629.

33. Buder, E.H., Vibratory Regime Classification of Infant Phonation. Journal of Voice, 2008. 22: p. 553–564.

34. Thelen, E. and Smith, L.B., A dynamic systems approach to the development of cognition and action. 1994, Cambridge: MIT Press.

35. Morgan, L.W., Y.E., A Systematic Review of the Literature on Early Vocalizations and Babbling Patterns in Young Children. Communication Disorders Quarterly, 2018. 40(1): p. 3–14.

36. Vihman, M.M., Phonological Development: The Origins of Language in the Child.. Vol. 312. 1996, Cambridge: Blackwell Publishers.

37. Masataka, N., Why early linguistic milestones are delayed in children with Williams syndrome: Late onset of hand banging as a possible rate-limiting constraint on the emergence of canonical babbling.. Developmental Science, 2001. 4(2): p. 158–164.

38. Oller, D.K. and Eilers, R.E., The role of audition in infant babbling. Child Development, 1988. 59: p. 441–449.

39. Eilers, R.E. and Oller, D.K., Infant vocalizations and the early diagnosis of severe hearing impairment. Journal of Pediatrics, 1994. 124: p. 199–203.

40. Vinter, S., L’analyse du babillage: une contribution au diagnostic de surdite?. Approche Neuropsychologique des Apprentissages chez l’Enfant, 1994. 6(4): p. 232–238.

41. Lynch, M.P., et al., The onset of speech-like vocalizations in infants with Down syndrome. American Journal of Mental Retardation, 1995. 100(1): p. 68–86.

42. Chapman, K., et al., Vocal development of 9 month-old babies with cleft palate.. Journal of Speech, Language and Hearing Research, 2001. 44: p. 1268–1283.

43. Lohmander, A., et al., Canonical Babbling and Early Consonant Development Related to Hearing in Children With Otitis Media With Effusion With or Without Cleft Palate. Cleft Palate Craniofac J, 2020: p. 1055665620966198.

44. Levin, K., Babbling in infants with cerebral palsy.. Clinical Linguistics & Phonetics, 1999. 13(4): p. 249–267.

45. Belardi, K.M., et al., A Retrospective Video Analysis of Canonical Babbling and Volubility in Infants with Fragile X Syndrome at 9-12 Months of Age. Journal of Autism and Developmental Disabilities, 2017.

46. Overby, M., K. Belardi, and J. Schreiber, A retrospective video analysis of canonical babbling and volubility in infants later diagnosed with childhood apraxia of speech. Clin Linguist Phon, 2020. 34(7): p. 634–651.

47. Patten, E., et al., Vocal patterns in infants with Autism Spectrum Disorder: Canonical babbling status and vocalization frequency. Journal of Autism and Developmental Disabilities, 2014. 44: p. 2413–2428.

48. Nathani, S., D.K. Oller, and A.R. Neal, On the robustness of vocal development: an examination of infants with moderate-to-severe hearing loss and additional risk factors. J Speech Lang Hear Res, 2007. 50(6): p. 1425–44.

49. Nyman, A., & Lohmander, A., Babbling in children with neurodevelopmental disability and validity of a simplified way of measuring canonical babbling ratio.. Clinical Linguistics & Phonetics, 2018. 32(2): p. 114–127.

50. Schramm, B., A. Bohnert, and A. Keilmann, The prelexical development in children implanted by 16 months compared with normal hearing children. Int J Pediatr Otorhinolaryngol, 2009. 73(12): p. 1673–81.

51. Molemans, I., et al., How to measure the onset of babbling reliably. Journal of Child Language, 2012. 39(3): p. 523–552.

52. Lieberman, M. and A. Lohmander, Observation is a valid way of assessing common variables in typical babbling and identifies infants who need further support. Acta Paediatr, 2014. 103(12): p. 1251–7.

53. Lang, S., et al., Canonical Babbling: A Marker for Earlier Identification of Late Detected Developmental Disorders? Curr Dev Disord Rep, 2019. 6(3): p. 111–118.

54. D. Xu, U.Y. S. Gray, Reliability of the LENA Language Environment Analysis System in young children’s natural home environment.. 2009, LENA Foundation: Boulder, CO. p. 1–16.

55. Oller, D.K., et al., Automated Vocal Analysis of Naturalistic Recordings from Children with Autism, Language Delay and Typical Development. Proceedings of the National Academy of Sciences, 2010. 107(30): p. 13354–13359.

56. Gilkerson, J., Richards, J. A., Warren, S. F., Oller, D. K., Russo, R., & Vohr, B. R., Language Experience in the Second Year of Life and Language Outcomes in Late Childhood. Pediatrics, 2018. 142(4): p. 1–11.

57. Warlaumont, A.S., et al., A social feedback loop for speech development and its reduction in autism. Psychol Sci, 2014. 25(7): p. 1314–24.

58. Yoder, P.J., Warren, S. F., & McCathren, R., Determining spoken language prognosis in children with developmental disabilities. American Journal of Speech-Language Pathology, 1998. 7: p. 77–87.

59. Warren, S.F., Gilkerson, J., Richards, J.A., & Oller, D.K., What Automated Vocal Analysis Reveals About the Language Environment of Young Children with Autism. Journal of Autism and Developmental Disabilities, 2010. 40: p. 555–569.

60. Koopmans-van Beinum, F.J.a.v.d.S. J.M., Early stages in the development of speech movements., in Precursors of early speech, B.L.a.R. Zetterstrom, Editor. 1986, Stockton Press: New York. p. 37–50.

61. Lewedag, V.L., Patterns of onset of canonical babbling among typically developing infants.. 1995, University of Miami: Coral Gables, FL.

62. Iyer, S.N. and D.J. Ertmer, Relationships between vocalization forms and functions in infancy: preliminary implications for early communicative assessment and intervention. Am J Speech Lang Pathol, 2014. 23(4): p. 587–98.

63. Törölä, H., Lehtihalmesa, M., Heikkinenc, H., Olsénb, P., & Ylihervaa, A., Early vocalization of preterm infants with extremely low birth weight (ELBW), part II: from canonical babbling up to the appearance of the first word. Clinical Linguistics & Phonetics, 2012. 26(4): p. 345–356.

64. Eilers, R.E., Oller, D.K., Levine, S., Basinger, D., Lynch, M.P. & Urbano, R., The role of prematuriy and socioeconomic status in the onset of canonical babbling in infants.. Infant Behavior and Development, 1993. 15: p. 297–315.

65. Oller, D.K., Eilers, R.E., Steffens, M.L., Lynch, M.P., Urbano, R., Speech-like vocalizations in infancy: an evaluation of potential risk factors. Journal of Child Language, 1994. 21(1): p. 33–58.

66. Oller, D.K., et al., Development of precursors to speech in infants exposed to two languages. J Child Lang, 1997. 24(2): p. 407–25.

67. Oller, D.K., et al., Infant boys are more vocal than infant girls. Curr Biol, 2020. 30(10): p. R426–R427.

68. Sahin, M. Early Biomarkers of Autism in Infants with TSC. 2013 [cited 2020 10/22/2020]; Available from: https://clinicaltrials.gov/ct2/show/NCT01780441.

69. Mundy, P.D. Christine & Goldstein, Jessica & Parlade, Meaghan & Hogan, Anne & Seibert, Jeffrey & Mundy, Dr., Early social communication scales (ESCS). 2003.

70. Bryson, S.E., et al., The Autism Observation Scale for Infants: scale development and reliability data. J Autism Dev Disord, 2008. 38(4): p. 731–8.

71. Mullen, E.M., Mullen Scales of Early Learning (AGS ed.). 1995, Circle Pines: American Guidance Service Inc.

72. Carter, A.S., et al., The Vineland Adaptive Behavior Scales: supplementary norms for individuals with autism. J Autism Dev Disord, 1998. 28(4): p. 287–302.

73. Sahli, A.S. and E. Belgin, Adaptation, validity, and reliability of the Preschool Language Scale-Fifth Edition (PLS-5) in the Turkish context: The Turkish Preschool Language Scale-5 (TPLS-5). Int J Pediatr Otorhinolaryngol, 2017. 98: p. 143–149.

74. Lord, C., et al., Autism diagnostic observation schedule: a standardized observation of communicative and social behavior. J Autism Dev Disord, 1989. 19(2): p. 185–212.

75. Lord, C., et al., Diagnosing autism: analyses of data from the Autism Diagnostic Interview. J Autism Dev Disord, 1997. 27(5): p. 501–17.

76. Delgado, R.E., E.H. Buder, and D.K. Oller AACT (Action Analysis Coding and Training). 2010, Miami: Intelligent Hearing Systems.

77. Oller, D.K., et al., Functional flexibility of infant vocalization and the emergence of language. Proc Natl Acad Sci U S A, 2013. 110(16): p. 6318–23.

78. Zimmerman, F., et al., Teaching By Listening: The Importance of Adult-Child Conversations to Language Development. Pediatrics, 2009. 124: p. 342–349.

